# Factors associated with suffering domestic violence in women during the last semester of the pandemic in Honduras- Central America

**DOI:** 10.1101/2023.02.04.23285439

**Authors:** Eleonora Espinoza-Turcios, Lysien Ivania Zambrano, Carlos Sosa-Mendoza, Henry Noel Castro Ramos, Dennis Arias-Chávez, Christian R. Mejia

## Abstract

**Introduction:** Mental health deterioration had already been reported prior to the pandemic, resulting in domestic violence in women, but this has not yet been reported in the pandemic in Central America.

**AIM:** To determine the Factors associated with suffering domestic violence in women in the last half year of the pandemic in Honduras.

**Methodology:** Analytical and retrospective cross-sectional study, carried out through a survey in 17 departments of Honduras, in hospitals and first level health care centers. The main variable was obtained through the question “if you suffered domestic violence in the last 6 months”, being the possible answer verbal, physical, psychological. Descriptive and analytical results were obtained.

**Results:** Of the 8442 Hondurans surveyed, 4.2% (352) perceived verbal violence, 1.9% (165) physical violence and 1.3% (113) psychological violence. In the multivariate analysis, it was found that women (p=0.001), those with a history of alcohol (p=0.002) or drug use (p=0.015), previous mental illness (p<0.001), mild (p<0.001), moderate (p<0.001) or severe (p=0.025) depression (p<0.001) had experienced more domestic violence; On the contrary, there was less perception of domestic violence at higher economic income (all p-values were <0.029), among single (p=0.003) and married people (p<0.001).

**Discussion:** Important Factors associated with suffering domestic violence in women, especially social factors and a history of mental illness, as well as alcohol and drug use in the home.

## INTRODUCTION

Domestic violence (DV) is defined as “a pattern of behavior that is used to gain or maintain power and control over an intimate partner in a relationship, may include a child or other relative, or any other member of the household” [1]. Globally, the lifetime prevalence of intimate partner violence is 26% among married or intimate partner women over the age of 15; estimating that 6% of women over the age of 15 have been subjected to non-partner sexual violence during some point in their lifetime [2].

At the beginning of the COVID-19 pandemic, in the first months of 2020, domestic violence increased worldwide, Spain recorded a 20% increase in calls to women’s helplines; [3]. United Kingdom reported an additional 25% one week after the announcement of the social distancing measures; [4] in France the increase of 36% was reported; [5] in Jingzhou-China the number of reported domestic violence cases was three times higher in February 2020, this compared to the same period last year [6]. This is in addition to the global and public health problems that were known prior to this pandemic that was generated worldwide, where one in three women had suffered in her life some type of physical and/or sexual violence by an intimate partner [7].

In Honduras, the problem of domestic violence behaves similarly to the rest of the world, and worsened as a result of the COVID-19 pandemic. According to data from the National Demographic and Health Survey (ENDESA /MICS 2019), 20% of Honduran women aged 15-49 years have ever been beaten or physically abused by someone after the age of 15, psychological violence occurs more frequently in urban areas (with 17% compared to 12% of cases in rural areas) and 4% of women aged 15-49 have suffered sexual abuse since the age of 12, being in 12.5% of cases produced by the current boyfriend/ex-boyfriend [8]. And as if this were not enough, according to the University Institute for Democracy, Peace and Security UNAH, there were 636 and 330 deaths of women due to violence in 2013 and 2021, respectively. And from January to October 2022 there were 252 violent deaths of women and/or femicide, that is, every 28 hours and 57 minutes a woman dies in Honduras [9]. The domestic violence women in Honduras involves multiple factors, such as socioeconomic, cultural, religious, and represents a serious social problem with serious consequences [10]. Study conducted by the National Autonomous University of Honduras through the Faculty of Medical Sciences ICIMEDES, that women suffer some type of violence 45% in five very low-income communities [11], also the national survey of the country ENDESA in 2019 shows that women between 15-49 years have suffered 16% some type of violence [8]. For all these reasons, the objective was to determine the Factors associated with suffering domestic violence in women in the last half year of the pandemic in Honduras.

## METHODOLOGY

An analytical, cross-sectional and retrospective research was generated. It was based on the information collected during the pandemic stage, which included the last third of the year 2021 and the first third of 2022. For the collection of information, a baseline instrument was elaborated, which measured the baseline mental health situation of the Honduran population in 17 of the departments [12]. This was carried out in health care centers, both hospitals and first level health care facilities.

In order for a respondent to participate in the research, it was taken into account that he/she was 18 years of age or older, that he/she agreed to participate in the research and that he/she had answered the main question, (“if you suffered domestic violence in the last 6 months”, being the possible answer verbal, physical, psychological). There were no exclusions because all respondents had answered the main question and met the other inclusion criteria. To determine whether the number of respondents was adequate for the main crosstabs, statistical power was calculated, where there was adequate power for the crosstabs of general perception of violence versus sex (power = 100%), according to the amount of family income (power = 100% for all categories), marital status (power = 100% for all categories), alcohol consumption in the last 6 months (power = 98%), drug consumption in the last 6 months (power = 100%), history of mental illness (power = 100%) and versus depression (power = 100% for all categories).

The main variable was obtained through the question “if you have suffered domestic violence in the last 6 months”, which was a semi-open question with one of four possible alternatives: no violence, or verbal, physical, psychological violence. For the individual analysis of each of the types of violence, they were crossed with each of the independent variables. But for the final analysis, the perception of violence was considered if one or more of the three categories (verbal, physical or psychological) was perceived, which was also crossed against the other variables. Secondary variables were age (in years completed), sex (male or female), amount of family income (<$400, $400- 800, $801-1199, $1200-1600 and >$1600), marital status (common-law/cohabiting, single, married or widowed/divorced), alcohol use in the past 6 months (no or yes), drug use in the past 6 months (no or yes), history of mental illness (no or yes) and versus depression (have not, mild, moderate, severe or very severe).

To proceed with the data analysis, the information was first extracted from the database, where an author did the general filtering of the data to be analyzed. Then a second quality control was carried out by another of the authors, where the variables were categorized, labeled and a glossary was constructed. Then all this information was passed to the statistical program Stata (license obtained by the authors who carried out the analysis). Then a table was created showing the frequencies and percentages of the suffering of the three types of domestic violence, then three tables were generated where these types of violence were crossed according to the socio-pathological variables, where the p-values were obtained with the Wilcoxon test (for the age variable) and the chi-square (for the other variables). Then a final table was generated, where the dependent variable was having suffered at least one type of violence, also being crossed against the socio-pathological variables; in this step the prevalence ratios, 95% confidence intervals and p-values were obtained, this with the use of generalized linear models: Poisson family, log link function and adjusted models for obtaining robust variances). The p-value <0.05 was the criterion for a variable to pass from the bivariate to the multivariate model, as well as to determine statistical significance.

Ethics were always respected, the initial study was approved by a local ethics committee (2019-62), The survey was anonymous, and participants were informed of the objective of the study.

## RESULTS

Of the 8442 Hondurans surveyed, 4.2% (352) perceived verbal violence, 1.9% (165) perceived physical violence and 1.3% (113) perceived psychological violence. When the 3 types of perceptions were added together, 97.9% (8265) did not perceive any violence, while 0.1% of respondents perceived all 3 types of violence in their homes during the last six months. Table **1**

**Table 1.** Frequencies and percentages of experiencing domestic violence in women during the last semester of the pandemic in Honduras, n=8442.

| Type of violence | Frequency | Percentage |
| --- | --- | --- |
| **Verbal abuse** |  |  |
| No | 8090 | 95.8 |
| Yes | 352 | 4.2 |
| **Physical violence\*** |  |  |
| No | 8277 | 98.1 |
| Yes | 165 | 1.9 |
| **Psychological violence** |  |  |
| No | 8329 | 98.7 |
| Yes | 113 | 1.3 |
| **Sum of perceptions** |  |  |
| No violence of any kind | 8265 | 97.9 |
| 1 type of violence | 126 | 1.5 |
| 2 type of violence | 45 | 0.5 |
| 3 type of violence | 6 | 0.1 |

\*don't include violence sexual.

Verbal domestic violence was associated with gender (p=0.009), family income (p<0.001), marital status (p=0.013), alcohol consumption at home (p<0.001) or drug use (p<0.001), a history of mental illness (p<0.001) and depression (p<0.001). Table 2

**Table 2.**
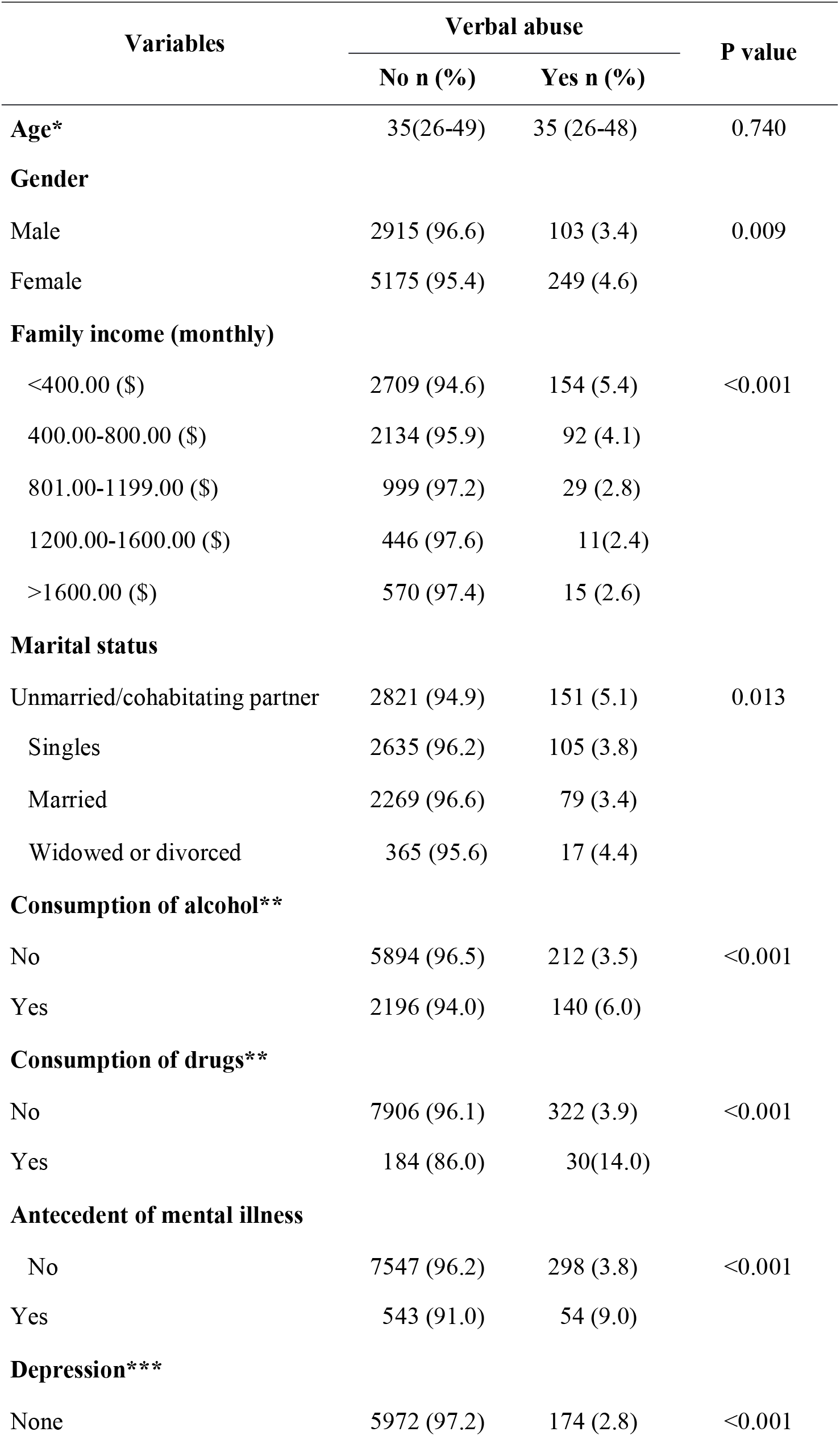
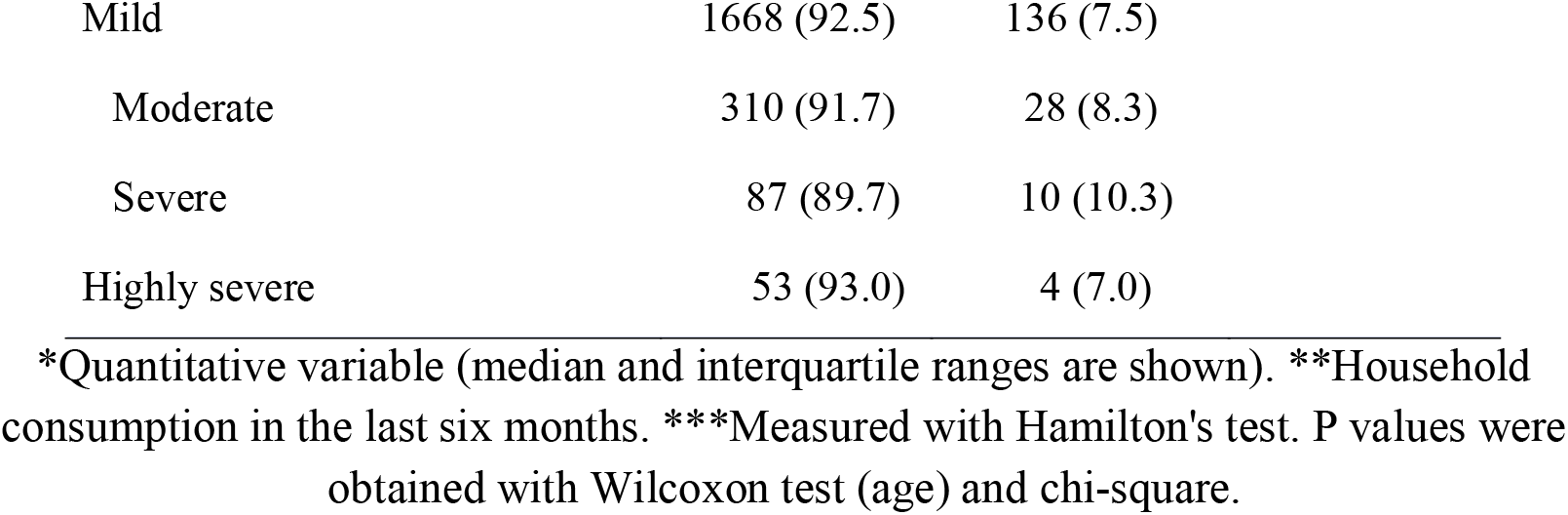
Household verbal abuse associated with socio-pathological variables in the last half year of the pandemic in Honduras, n=8442.

| Variables | Verbal abuse |  | P value |
| --- | --- | --- | --- |
|  | No n (%) | Yes n (%) |  |
| **Age\*** | 35(26-49) | 35 (26-48) | 0.740 |
| **Gender** |  |  |  |
| Male | 2915 (96.6) | 103 (3.4) | 0.009 |
| Female | 5175 (95.4) | 249 (4.6) |  |
| **Family income (monthly)** |  |  |  |
| <400.00 (\$) | 2709 (94.6) | 154 (5.4) | <0.001 |
| 400.00-800.00 (\$) | 2134 (95.9) | 92 (4.1) | |
| 801.00-1199.00 (\$) | 999 (97.2) | 29 (2.8) | |
| 1200.00-1600.00 (\$) | 446 (97.6) | 11(2.4) | |
| >1600.00 (\$) | 570 (97.4) | 15 (2.6) | |
| **Marital status** |  |  |  |
| Unmarried/cohabitating partner | 2821 (94.9) | 151 (5.1) | 0.013 |
| Singles | 2635 (96.2) | 105 (3.8) |  |
| Married | 2269 (96.6) | 79 (3.4) |  |
| Widowed or divorced | 365 (95.6) | 17 (4.4) |  |
| **Consumption of alcohol\*\*** |  |  |  |
| No | 5894 (96.5) | 212 (3.5) | <0.001 |
| Yes | 2196 (94.0) | 140 (6.0) |  |
| **Consumption of drugs\*\*** |  |  |  |
| No | 7906 (96.1) | 322 (3.9) | <0.001 |
| Yes | 184 (86.0) | 30(14.0) |  |
| **Antecedent of mental illness** |  |  |  |
| No | 7547 (96.2) | 298 (3.8) | <0.001 |
| Yes | 543 (91.0) | 54 (9.0) |  |
| **Depression\*\*\*** |  |  |  |
| None | 5972 (97.2) | 174 (2.8) | <0.001 |

|  |  |  |
| --- | --- | --- |
| Mild | 1668 (92.5) | 136 (7.5) |
| Moderate | 310 (91.7) | 28 (8.3) |
| Severe | 87 (89.7) | 10 (10.3) |
| Highly severe | 53 (93.0) | 4 (7.0) |

\*Quantitative variable (median and interquartile ranges are shown). \*\*Household consumption in the last six months. \*\*\*Measured with Hamilton's test. P values were obtained with Wilcoxon test (age) and chi-square.

Associated with physical violence at home were home use of alcohol (p=0.004) or drugs (p<0.001), history of mental illness (p<0.001) and depression (p<0.001). Table 3

**Table 3.**
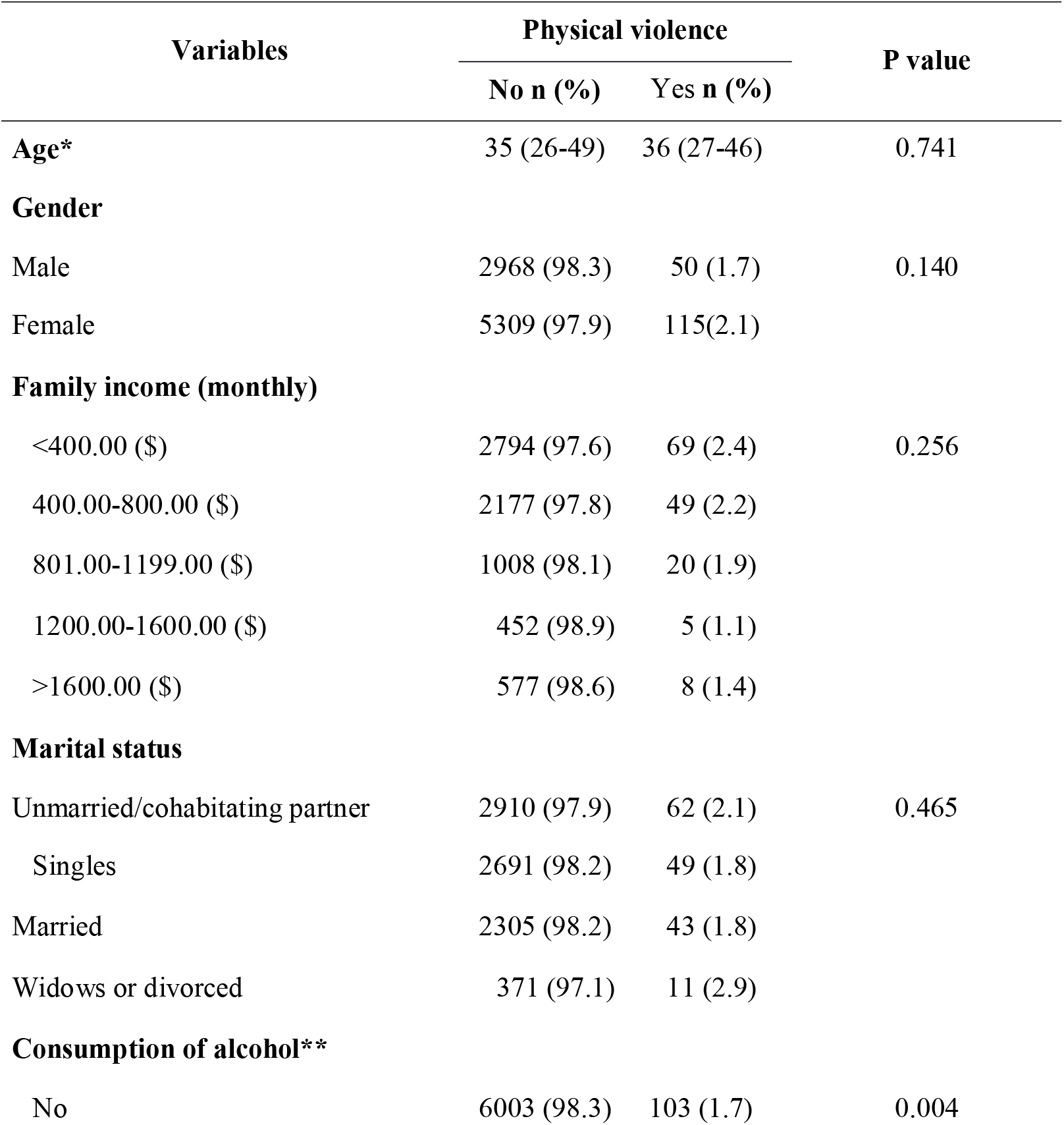
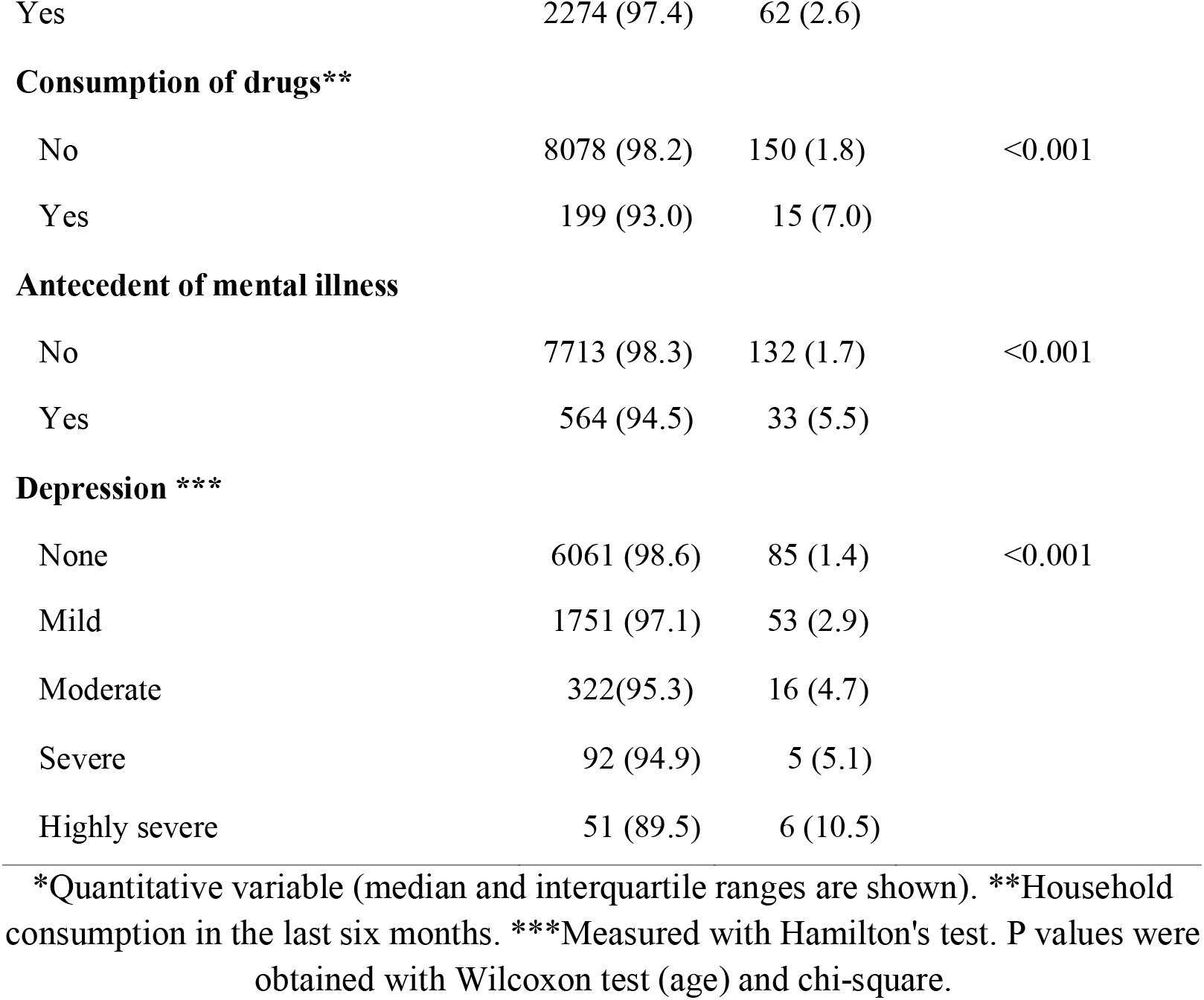
Physical violence at home associated with socio-pathological variables in the last half year of the pandemic in Honduras, n=8442.

Family income (p=0.031), home consumption of alcohol (p<0.001) or drugs (p<0.001), history of mental illness (p<0.001) and depression (p<0.001) were associated with verbal violence at home. Table 4

**Table 4.** Domiciliary sexual violence associated with socio-pathological variables in the last half year of the pandemic in Honduras, n=8442.

| Variables | Sexual violence |  | P value |
| --- | --- | --- | --- |
|  | No n (%) | Si n (%) |  |
| **Age** | 35 (26-49) | 36 (29-47) | 0.808 |
| **Gender** |  |  |  |
| Male | 2985 (98.9) | 33 (1.1) | 0.144 |
| Female | 5344 (98.5) | 80 (1.5) |  |
| **Family income (monthly)** |  |  |  |
| <400.00 (\$) | 2810 (98.2) | 53 (1.8) | 0.031 |
| 400.00-800.00 (\$) | 2200 (98.8) | 26 (1.2) | |
| 801.00-1199.00 (\$) | 1017 (98.9) | 11 (1.1) | |
| 1200.00-1600.00 (\$) | 456 (99.8) | 1 (0.2) | |
| >1600.00 (\$) | 576 (98.5) | 9 (1.5) | |
| **Marital status** |  |  |  |
| Unmarried/cohabitating partner | 2927 (98.5) | 45 (1.5) | 0.554 |
| Singles | 2710 (98.9) | 30 (1.1) |  |
| Married | 2316 (98.6) | 32 (1.4) |  |
| Widows or divorced | 376 (98.4) | 6 (1.6) |  |
| **Consumption of alcohol\*\*** |  |  |  |
| No | 6042 (99.0) | 64 (1.0) | <0.001 |
| Yes | 2287 (97.9) | 49 (2.1) |  |
| **Consumption of drugs\*\*** |  |  |  |
| No | 8124 (98.7) | 104 (1.3) | <0.001 |
| Yes | 205 (95.8) | 9 (4.2) |  |
| **Antecedent of mental illness** |  |  |  |
| No | 7760 (98.9) | 85 (1.1) | <0.001 |
| Yes | 569 (95.3) | 28 (4.7) |  |
| **Depression\*\*\*** |  |  |  |
| None | 6098 (99.2) | 48 (0.8) | <0.001 |
| Mild | 1753 (97.2) | 51 (2.8) |  |
| Moderate | 328 (97.0) | 10 (3.0) |  |
| Severe | 95 (97.9) | 2 (2.1) |  |
| Highly severe | 55 (96.5) | 2 (3.5) |  |

\*Quantitative variable (median and interquartile ranges are shown). \*\*Household consumption in the last six months. \*\*\*Measured with Hamilton's test. P values were obtained with Wilcoxon test (age) and chi-square.

In the multivariate analysis, it was found that women (PRa: 1.92; 95%CI: 1.33- 2.77; p-value=0.001), those with a history of alcohol (PRa: 1.77; 95%CI: 1.24-2.52; p- value=0.002) or drug use (PRa: 2.08; 95%CI: 1.15-3.75; p-value=0.015), previous mental illness (PRa: 2.94; 95%CI: 2.00-4.31; p-value<0.001), mild (RPa: 2.48; 95%CI: 1.75-3.53; p-value<0.001), moderate (RPa: 3.24; 95%CI: 1.85-5.70; p-value<0.001) or severe depression (RPa: 3.11; 95%CI: 1.15-8.39; p-value=0.025); on the contrary, there was less perception of domestic violence at higher economic income (all p-values were <0.029), among single (p=0.003) and married (p<0.001). **Table 5**

**Table 5.** Bivariate and multivariate analysis of factors associated domestic violence in women during the last semester of the pandemic in Honduras, n=8442.

| Variable | Bivariate analysis | Multivariate analysis |
| --- | --- | --- |
| **Age\*** | 0.998 (0.988-1.008) 0.685 | Not entered in the final model |
| **Gender female** | 1.68 (1.20-2.36) 0.003 | 1.92 (1.33-2.77) 0.001 |
| **Family income (monthly)** |  |  |
| <400.00 (\$) | Comparison category | Comparison category |
| 400.00-800.00 (\$) | 0.63 (0.44-0.90) 0.012 | 0.65 (0.45-0.93) 0.018 |
| 801.00-1199.00 (\$) | 0.39 (0.21-0.71) 0.002 | 0.40 (0.21-0.75) 0.004 |
| 1200.00-1600.00 (\$) | 0.36 (0.15-0.89) 0.027 | 0.36 (0.14-0.89) 0.028 |
| >1600.00 (\$) | 0.34 (0.15-0.78) 0.010 | 0.30 (0.13-0.68) 0.004 |
| **Marital status** |  |  |
| Unmarried/cohabitating partner | Comparison category | Comparison category |
| Singles | 0.51 (0.36-0.72) <0.001 | 0.56 (0.38-0.82) 0.003 |
| Married | 0.37 (0.24-0.56) <0.001 | 0.43 (0.27-0.68) <0.001 |
| Widows or divorced | 0.65 (0.32-1.32) 0.234 | 0.56 (0.25-1.26) 0.160 |
| **Consumption of alcohol\*\*** | 1.52 (1.12-2.05) 0.007 | 1.77 (1.24-2.52) 0.002 |
| **Consumption of drugs\*\*** | 3.05 (1.76-5.27) <0.001 | 2.08 (1.15-3.75) 0.015 |
| **Antecedent of mental illness** | 3.71 (2.63-5.25) <0.001 | 2.94 (2.00-4.31) <0.001 |
| **Depression\*\*\*** |  |  |
| None | Comparison category | Comparison category |
| Mild | 2.92 (2.14-3.98) <0.001 | 2.48 (1.75-3.53) <0.001 |
| Moderate | 3.03 (1.73-5.28) <0.001 | 3.24 (1.85-5.70) <0.001 |
| Severe | 3.77 (1.56-9.09) 0.003 | 3.11 (1.15-8.39) 0.025 |
| Highly severe | 2.57 (0.65-10.18) 0.180 | 1.40 (0.22-8.70) 0.721 |

\*Quantitative variable. \*\*Household consumption in the last six months. \*\*\*Measured with Hamilton's test. Prevalence ratios are shown to the left of the parentheses, 95% confidence intervals within the parentheses and p-values to the right (all obtained with generalized linear models: Poisson family, log link function and models adjusted for robust variances).

## DISCUSSION

The objective of this study was to determine the factors associated with suffering domestic violence in women during the pandemic in Honduras, since, Honduras does not have updated national studies, the 2017 Chavarria-Mejia study was conducted in two communities in Honduras, where they found that 79% of the 153 women over 16 years of age with a partner, had some type of domestic violence and 40% presented economic violence [10].

In another study on domestic violence in women in five communities in Honduras, the overall prevalence was 45% (737), the main type of violence was verbal 52% (383), but psychological violence was also important, 4% (31), sexual violence 19% (140) and physical violence 21% (151), predominantly in women between 20-39 years of age [11].

The 2019 ENDESA reported among women aged 15-49 years that 16% have experienced some type of violence (psychological, physical or sexual) by a husband/partner, 15% have suffered psychological violence, 6%, physical violence and 2% sexual violence [8]. This is aggravated by the report that practically once a day a woman dies in this country [9].

As can be seen in the three previous studies, the report of violence has been obtained in different populations in Honduras, but above all among women and prior to the COVID-19 pandemic, therefore, this was obtained in a population that attends health facilities, which are generally those who have more access to health care and have other underlying diseases. In addition, the results we present are important to be able to have a current statistic and that is given after one of the most important events for public health in recent decades [13].

The latter being very important because during the confinement that generated COVID-19, domestic violence increased due to the continuous contact between perpetrators and victims; an increase in cases has been reported in North America, Europe, Asia, the Pacific and Africa, especially during the first week of the blockade in each country [14]. A systematic review showed an increase in domestic violence following lockdowns [15].

As a first important result, less than 5% of the respondents perceived some type of violence. These results are similar to those described in Germany, where an online survey of 3545 volunteers reported that 5% of all participants (5.1% of all women and 4.1% of all men) reported having experienced interpersonal violence (IPV), verbal (98% of all women and 100% of all men who experienced IPV), physical (38.4% of all women and 63.6% of all men who experienced IPV), or sexual (26.5% of all women and 50% of all men who experienced IPV) [16]. This is relevant because of the large number of people affected globally, as calculated in the United States of America, and it affects the victim, families, co-workers and the community, causes a decrease in psychological and physical health, a decrease in quality of life and a decrease in productivity [17]. Therefore, these results should be considered exploratory, since the objective was not to extrapolate the data to the entire Honduran population, but they are important because they were taken from a large population in the most important departments of the country and at a time when the pandemic was still present among the respondents.

Another important result was that women had a higher frequency of perceived domestic violence. Knowing that there are global reports indicating that an estimated 243 million women and adolescents aged 15-49 years have been victims of physical and/or sexual violence at the hands of an intimate partner [18]. A study conducted in 2020 in the United States reported that the prevalence of domestic violence during the COVID-19 pandemic was 18%; and that changes in income and employment increased the incidence of domestic violence by 1.63 times [19]. In another study, also from 2020 in the USA, it was reported that the incidence of physical domestic violence against women was 1.8 times higher than before [20].

This is in addition to other important results that have been reported in other realities, such as in Peru in 2020 where a 48% increase in the number of calls made to the Domestic Violence Control and Assistance Unit was reported [21]. A meta-analysis showing an increase in psychological/emotional as well as verbal violence in the general population during the COVID-19 pandemic [22]. The World Vision report on domestic violence hotline calls during the outbreak of the COVID-19 pandemic shows that there was an increase in domestic violence cases worldwide; more specifically, in Argentina (25%), Bosnia and Herzegovina (22%), Brazil (18%), Chile (75%), Cyprus (47%), France (30%), India (32%); Lebanon (50%), Mexico (25%), Montenegro (27%), Singapore (33%), Spain (12%) and in the United States (22%); which were mainly related to intimate partner violence [23].

In South Africa, it was found that the fear and stress associated with pandemics can exacerbate intimate partner violence through control, manipulation and coercion tactics, as well as confinement measures that give abusers greater freedom to act without inspection or consequences [24]. Latin America and the Caribbean experienced increases in unemployment, poverty, food insecurity, etc. [25]. The prevalence of mental health symptoms during the COVID-19 crisis in Latin America reported that anxiety levels were significantly higher than in other regions, such as China (25%; p < 0.001) [26]. There was also an increase in reported cases of domestic violence, particularly toward child abuse and intimate partner violence against women [27]. Exacerbating the high rates of violence in the region, which were reported to be three times the global average before the pandemic [28]. And at the national level, Honduras lives in a normalized environment of violence, from the political violence that the country suffers, which is also influenced by living in a conservative environment and finding 60% of low educational level and 8% with no education; where it can be assumed that violence may not be identified or women are not allowed to talk about it [29]. All these cases give us a picture of how we can be in an environment that generates greater abuse for women and other vulnerable populations, which should be evaluated with greater emphasis in future research.

We also found that the higher the level of family income, the less domestic violence there was. Kebede et al. analyzed Demographic and Health Survey data from 187716 women with partners aged 10-59 years from 20 low-and middle-income countries; where the findings suggest that the relationship between wealth and violence varies considerably in low- and middle-income countries [30]. This is in addition to the fact that the economic and labor crisis created by the pandemic could increase global unemployment by almost 25 million, according to a new assessment by the International Labor Organization (ILO) [31]. On the other hand, during the time of confinement, an increasing number of poor families do not have a source of daily income, which generates frustration and feelings of helplessness, all of which creates conditions for increased violence [32].

This is reinforced by the finding that a structured questionnaire on intimate partner violence found that wealthier and more empowered women reported less intimate partner violence, and that younger women and those living in rural areas tended to be more exposed to intimate partner violence [33]. That, if added to the other factors that were generated by movement restrictions, increases in child abuse, internet addiction, domestic violence and divorce consultations, as well as suicide rates [34]. All of this together tells us of a large social component that is closely linked to the whole picture of violence, which should be addressed in more detail by sociologists, psychologists, and other professionals.

We also found in this study that the consumption of alcohol or drugs in the home was associated with a higher frequency of domestic violence, it should be noted that the relationship between the consumption of alcohol and psychoactive substances with domestic violence is already well established [35, 36]. Having found clear evidence of a temporal association between episodes of alcohol consumption and subsequent episodes of occurrence of verbal and physical aggression [37]. In addition, the acceptance of alcohol consumption by men normalizes consumption and amplifies other behaviors tolerated in macho cultures, such as the use of violence [38]. This may also be due to the accumulation of stressful events, which, in the context of the current pandemic was precipitated by isolation, which favored the intensification of alcohol consumption within the family environment [39]. A systematic review reported that frequent alcohol consumption by men increased the risk of perpetration of violence [40]. Another meta- analysis showed that people at risk for domestic violence and substance abuse are chronically exposed to elevated levels of anxiety, highly stressful environments, and unfavorable economic situations [41]. Because of all this evidence, it is important to screen families who may have alcohol, drug, or other problems that are closely related to domestic violence.

Our results reported that less domestic violence was found among single and married people. This is important to analyze within Honduran society, which, like other Latin American countries, has an important male chauvinist component, where gender roles influence individual behavior and relationships, due to the power and control that men try to impose on the entire family [42]. Therefore, what was reported is important as a starting point to determine the role that men, women and other members of the household play in the violence that may be perceived.

Yuan K, reported that women showed a higher prevalence of depression, anxiety and insomnia, women younger than 40 years or in the upper income level had a lower prevalence of depression and anxiety [43]. Having a history of mental illness was also associated with a higher frequency of domestic violence. COVID-19 may have a substantial impact on mental health and well-being. Jung et al showed that 25% of participants reported pre-existing mental health problems and showed more depression and anxiety, less well-being, less sense of coherence, and worse coping skills in terms of the pandemic and actions taken [16]. Thus, intimate partner violence against women is frequently and strongly associated with mental health problems [44]. Vai reported that female sex, having a prior psychiatric diagnosis, and psychopathology at 1-month follow-up were moderators of depression in the post-COVID-19 syndrome [45].

Finally, depression was associated with women suffering from domestic violence, Robinson 2022 reported that there was a small increase in mental health symptoms shortly after the outbreak of the COVID-19 pandemic in symptoms of depression and mood disorders, which tended to be higher and remained significantly elevated in May-July [46]. A systematic review found that suggested risk factors for violence against women and girls after disasters include increased life stressors, law enforcement failure, exposure to high-risk environments, exacerbation of existing gender inequalities, and unequal social norms [47]. Which is increased when there is domestic conflict and violence, financial loss, anxiety and depression, and pre-existing mental health conditions, all of which should be considered to prevent suicide attempts and deaths secondary to the COVID-19 pandemic [48].

### Limitations

The main limitation was selection bias, since the data obtained cannot be extrapolated to the entire Honduran population; however, by obtaining good statistical power, the associations and results can be used to generate important conclusions that can be used for the elaboration of research that aims to have more associated variables, with a larger sample size and representativeness, as well as with more advanced designs. In addition, there could have been a possible bias because some may have hidden or changed their answers out of fear or embarrassment; however, it is believed that, if it occurred, it was minimal, since the anonymous nature of the survey helped them to have the confidence to answer truthfully.

## Conclusions

From all the findings it is concluded that women had a higher frequency of domestic violence, the consumption of alcohol or drugs in the home, having a history of mental illnesses such as depression, likewise, there was less frequency of domestic violence according to a higher economic income in the home, between single and married people.

The inequality of opportunities, the responsibility of Honduran households that falls on women as heads of household, extreme poverty, and low schooling make women more vulnerable to domestic violence. Therefore, it is important to continue with this line of research in order to have real data at the national level and make decisions to address this problem.

## Data Availability

All data produced in the present work are contained in the manuscript

https://unahedu-my.sharepoint.com/:f:/r/personal/eleonora_espinoza_unah_edu_hn/Documents/Base%20de%20datos?csf=1&web=1&e=8GGF04

## Author Contributions

Study design: CM, EET, LIZ, and CASM Data collection: CM, HNCR Data analysis: CM, DAC Writing: CM, EET, LIZ, CASM, DAC, and HNCR, all authors have read and agreed to the published version of the manuscript.

## Institutional Review Board Statement

The study was conducted under the Declaration of Helsinki. This research’s preparation and execution fully complied with the fundamental ethical principles of autonomy, justice, beneficence, and non- maleficence. The Act Number (2019062), approved by the Ethics Committee in Biomedical Research (CEIB) of the National Autonomous University of Honduras (UNAH), meeting of December 02, 2021.

## Data Availability Statement

The data presented in this study are available on request from the corresponding author.

## Acknowledgments

To all MSS of the cohort who agreed to participate in the project, from September to March 2021-2022 and from October-April 2021-2022, to Mr. Mauricio Gonzales for the elaboration of the database and a first filtering of the database.

